# Central cholinergic degeneration in prodromal and early manifest Parkinsons disease: relation to cognition, clinical phenotype, and future conversion

**DOI:** 10.1101/2024.12.31.24319810

**Authors:** Tamir Eisenstein, Karolien Groenewald, Ludo van Hillegondsberg, Falah Al Hajraf, Tanja Zerenner, Michael A Lawton, Yoav Ben-Shlomo, Ludovica Griffanti, Michele T Hu, Johannes C Klein

**Affiliations:** Wellcome Centre for Integrative Neuroimaging, FMRIB, Nuffield Department of Clinical Neurosciences, University of Oxford; Oxford Parkinson’s Disease Centre, Nuffield Department of Clinical Neurosciences, University of Oxford, Oxford, UK; Department of Pharmacology & Toxicology, Faculty of Medicine, Kuwait University, Kuwait City, Kuwait; Population Health Sciences, Bristol Medical School, University of Bristol, Bristol, UK; Oxford Centre for Human Brain Activity, Wellcome Centre for Integrative Neuroimaging, Department of Psychiatry, University of Oxford

## Abstract

The neuropathological process in Parkinson’s disease (PD) and Lewy body disorders has been shown to extend well beyond the degeneration of the dopaminergic system, affecting other neuromodulatory systems in the brain which play crucial roles in the clinical expression and progression of these disorders.

Here, we investigate the role of the macrostructural integrity of the nucleus basalis of Meynert (NbM), the main source of cholinergic input to the cerebral cortex, in cognitive function, clinical manifestation, and disease progression in non-demented subjects with PD and individuals with isolated REM sleep behaviour disorder (iRBD).

Using structural MRI data from 393 early PD patients, 128 iRBD patients, and 186 controls from two longitudinal cohorts, we found significantly lower NbM grey matter volume in both PD (β=-12.56, *p*=0.003) and iRBD (β=-16.41, *p*=0.004) compared to controls. In PD, higher NbM volume was associated with better higher-order cognitive function (β=0.12, *p*=0.012), decreased non-motor (β=-0.66, *p*=0.026) and motor (β=-1.44, *p*=0.023) symptom burden, and lower risk of future conversion to dementia (Hazard ratio (HR)<0.400, *p*<0.004). Higher NbM volume in iRBD was associated with decreased future risk of phenoconversion to PD or dementia with Lewy bodies (DLB) (HR<0.490, *p*<0.016). However, despite similar NbM volume deficits to those seen in PD, associations between NbM structural deficits and current disease burden or clinical state were less pronounced in iRBD.

These findings identify NbM volume as a potential biomarker with dual utility: predicting cognitive decline and disease progression in early PD, while also serving as an early indicator of phenoconversion risk in prodromal disease. The presence of structural deficits before clear clinical correlates in iRBD suggests complex compensatory mechanisms may initially mask cholinergic dysfunction, with subsequent failure of these mechanisms potentially contributing to clinical conversion.

## Introduction

Parkinson’s disease (PD) is the second most prevalent neurodegenerative disease, affecting >1% of the population ≥65 years of age^1^. Despite being generally classed as a movement disorder, PD is associated with a spectrum of non-motor features which are key drivers of disease impact, especially in early PD^2–4^. Even in the absence of dementia, multiple cognitive domains are affected, in particular complex higher-order functions such as executive functions, attention, visuospatial skills, and memory^5^. Cognitive deficits have been shown to be up to 6 times more prevalent in PD compared to the general population, with PD-related dementia (PDD) arising in up to 80% of patients after two decades following diagnosis^6,7^. In addition, a large variability exists in time to dementia with some developing dementia within the first few years after diagnosis while others remain resilient to it for decades. However, despite being one of the most disabling non-motor manifestations of PD, there are still no robust predictors of dementia that are well validated. These would be valuable to enrich target populations for clinical trials aiming to prevent or delay cognitive deterioration^8^.

Neuronal loss in the substantia nigra, dopamine deficiency in the striatum, and intracellular aggregates of a-synuclein (aSyn), i.e., Lewy bodies, are considered the neuropathological hallmarks of PD and Lewy body disorders. However, Lewy body disorders extend well beyond the degeneration of the dopaminergic system, and other neuromodulatory systems in the brain play crucial roles in the clinical expression and progression of these disorders^9^. The basal forebrain cholinergic system (BFCS) is the major source of cholinergic innervation to the neocortex, hippocampus, and amygdala^10^, and is largely divided into two distinct subregions, namely Ch1-3 which includes the medial septum and diagonal band of Broca nuclei and Ch4 which includes the nucleus basalis of Meynert (NbM). BFCS neurons have been shown to provide important control over circuit dynamics underlying behavioural and cognitive processing, in particular attention, visuospatial skills, and memory^11,12^, which are cognitive domains affected in PD and other Lewy body disorders^5,13^.

Loss of the cholinergic innervation to the cerebral cortex has been suggested as a mechanism for cognitive decline and dementia in Lewy body disorders^5^, and the accumulation of aSyn deposition within the BFCS neurons has been suggested to occur simultaneously with neuronal loss in the substantia nigra^14^. Neuroimaging methods such as positron emission tomography (PET) and magnetic resonance imaging (MRI) have provided evidence for central cholinergic degeneration and dysfunction in Lewy body disorders with and without clinically significant cognitive impairment including PD, PDD, and dementia with Lewy bodies (DLB). Cholinergic dysfunction has been shown to be more prominent in DLB/PDD than PD^15–19^. Furthermore, not all subregions of the BFCS are equally susceptible to the neuropathological process of Lewy body disorders, and NbM neurons may be more vulnerable than other subparts of this system^16,20^. Therefore, the association between cognitive impairment and cholinergic deficits in Lewy body disorders and the high vulnerability of the NbM in these disorders make it a potential biomarker for prognostication among these patients.

Given that objective cognitive impairment is already prevalent in 10-20% of early PD patients^5,8^, and that PD diagnosis by itself constitutes a risk factor for dementia, there is a great need to identify patients at greater risk for PD at the prodromal stages of the disease if future targeted interventions are to be successful. Furthermore, given that cholinergic deficits are well associated with cognitive deficits in PD, they may also serve as potential disease biomarkers for cognitive progression in the prodromal phase.

PD is characterized by a notably long and diverse prodromal stage which could span decades^21,22^. Many of the prodromal markers of PD are not specific to Lewy body disorders. This includes depression, anxiety, olfactory loss, and autonomic change. A notable exception is isolated rapid eye movement (REM) sleep behaviour disorder (iRBD)^23^. iRBD is a sleep disorder characterized by the loss of muscle atonia during the REM sleep state, leading to dream enactment^24^, and considered one of the strongest markers for prodromal PD and aSyn aggregation neurodegenerative disorders. Individuals with iRBD are at high risk for a clinical diagnosis of manifest Lewy body disorder, and previous longitudinal multicenter studies found phenoconversion rates to aSyn neurodegeneration of 6-8% per year and >60% risk of phenoconversion after a decade^25–27^. In addition, among converters, roughly equal proportions progress to PD (parkinsonism first) or DLB (dementia first)^26^. Furthermore, concomitant RBD may occur in ∼33% of early PD patients within 3 years of diagnosis, and associates with higher baseline motor and cognitive dysfunction, as well as faster motor and non-motor progression^28^. However, as with the case of dementia among PD patients, time to phenoconversion in iRBD is highly variable and may occur years to even decades after the onset of RBD symptoms. While iRBD is characterized by prominent degeneration of brainstem and peripheral cholinergic nuclei^24^, the extent of central cholinergic changes that take place in the brain at this prodromal phase are less understood, and whether baseline BFCS deficits may serve as a potential marker of future phenoconversion in these patients is unclear.

Here, we combined two longitudinal cohorts of PD and iRBD patients, the Oxford Discovery Cohort (ODC) and the Parkinson’s Progression Markers Initiative (PPMI) to (1) examine whether macrostructural deficits in the NbM are already evident at the stage iRBD and among early PD patients without cognitive impairment; (2) whether reduced NbM grey matter volume is associated with worse cognitive and clinical measures at the baseline visit; and (3) whether lower NbM grey matter volume is associated with a higher risk of conversion from PD to PDD and (4) higher risk of phenoconversion to a definitive neurodegenerative disorder in iRBD.

## Materials and methods

### Participants

Data from a total of 393 PD patients within 3 years of diagnosis, 128 participants with iRBD, and 186 healthy controls were included in this study from the multisite PPMI database (https://www.ppmi-info.org) and the ODC, which is a prospective, longitudinal study that has recruited patients with early idiopathic Parkinson’s, healthy controls (HCs) and individuals at risk of PD since 2010^29,30^. See Supplementary Table 1 for the contribution of each study to the overall number of groups’ participants. PPMI and ODC are longitudinal observational studies with a comprehensive set of clinical and MRI measures^29,30^. Data were last checked for updates and downloaded in September 2024. Participants’ demographics are summarised in Table 1. iRBD patients in both cohorts were diagnosed with sleep laboratory-based polysomnography and were free of parkinsonism at recruitment. All participants underwent the MDS-Unified Parkinson’s Disease Rating Scale (UPDRS) to assess motor and non-motor symptoms, as well as the Montreal Cognitive Assessment (MoCA) to evaluate general cognitive functioning. Both cohort studies were approved by local ethics committees and all participants provided written informed consent according to the Declaration of Helsinki.

**Table 1.** demographic, cognitive, and clinical characteristics of study participants (mean±SD and proportions are presented for continuous and categorical variables, respectively).

| **Characteristic** | **PD**<br>**(N = 393)** | **iRBD**<br>**(N = 128)** | **HC**<br>**(N = 186)** |
| --- | --- | --- | --- |
| Age (years) | 63.5 (9.54) | 67.2 (7.23) | 62.6 (10.91) |
| Sex |  |  |  |
| Female | 149 (38%) | 13 (10%) | 68 (37%) |
| Male | 244 (62%) | 115 (90%) | 118 (63%) |
| Education (years) | 15.7 (3.12) | 14.4 (3.40) | 16.0 (3.09) |

| Characteristic | PD<br>(N = 393) | iRBD<br>(N = 128) | HC<br>(N = 186) |
| --- | --- | --- | --- |
| MoCA score | 27.3 (2.10) | 26.3 (2.30) | 27.9 (2.00) |
| Higher order cognition<br>(z-score) | -0.01<br>(0.64)<br>(N = 282) | 0.00 (0.68)<br>(N = 48) | - |
| UPDRS-1 total score | 6.4 (4.30) | 8.5 (5.13) |  |
| UPDRS-3 total score | 22.8 (9.68) | 4.1 (3.37) | - |
| Disease duration<br>(months) | 9.1 (8.96) | 27.3<br>(44.37) | - |
| TIV (ml) | 1,531.2<br>(152.86) | 1,541.1<br>(125.34) | 1,495.9<br>(146.81) |
| IQR (%) | 80.7 (3.66) | 81.6 (3.95) | 80.3 (3.74) |

### Magnetic resonance imaging (MRI)

Multisite data of 3T high resolution T1-weighted MRI scans (1 x 1 x 1 mm^3^) from the baseline visit of PPMI were obtained in compliance with the PPMI data agreement. Acquisition parameters and detailed protocols are described on the PPMI website (https://www.ppmi-info.org/study-design/research-documents-and-sops). Data acquisition in ODC was performed at the Oxford Centre for Clinical Magnetic Resonance Research (OCMR) using a 3T Siemens Trio MRI scanner equipped with a 12-channel coil. T1-weighted images were obtained using a 3D magnetization prepared-rapid acquisition gradient echo (MPRAGE) sequence (192 axial slices, flip angle 8°, 1 x 1 x 1 mm^3^ voxel size, echo time/repetition time/inversion time = 4.7 ms/2040 ms/900 ms)^30^.

### Grey matter (GM) volume assessment

Measurements of the NbM GM volume were derived from each participant’s T1-weighted MR image using the Computational Anatomy Toolbox (CAT12, https://neuro-jena.github.io/cat/)^31,32^ implemented in Statistical Parametric Mapping (SPM12) software. Default CAT12 preprocessing steps were used to process the raw T1-weighted images such as correcting for bias field inhomogeneities, affine-registration, and segmentation into brain tissue classes (i.e., grey matter (GM), white matter (WM), and cerebrospinal fluid (CSF)). Further preprocessing included local tissue intensity transformation, partial volume estimation, and spatial normalization to standard MNI space using DARTEL. The spatially normalized images were then “modulated” by multiplying the voxel values with the Jacobian determinant (i.e., linear and non-linear components) derived from the spatial normalization. This allows the extraction of the tissue volume (e.g. "concentration" of grey matter)^33^. The GM volumes of the right and left NbM were extracted from the modulated, normalized, partial volume-corrected GM images of each participant using Ch4 masks from the probabilistic cytoarchitectonic map of the Basal Forebrain (v4.2)^34,35^, in accordance with the suggested threshold for realistic NbM volume estimation^36^ (**Figure 1**). In addition to NbM volume, we extracted the total intracranial volume (TIV) of each participant to control for differences in head size. Images were visually inspected and excluded if image quality was not sufficient, or if the image quality rating (IQR) score was below 70%. The IQR is a composite measurement generated by the CAT12 pipeline, integrating several metrics of image quality into a single value ranging from 0 to 1 (i.e., the higher the score, the better the image quality).

**Figure 1.**
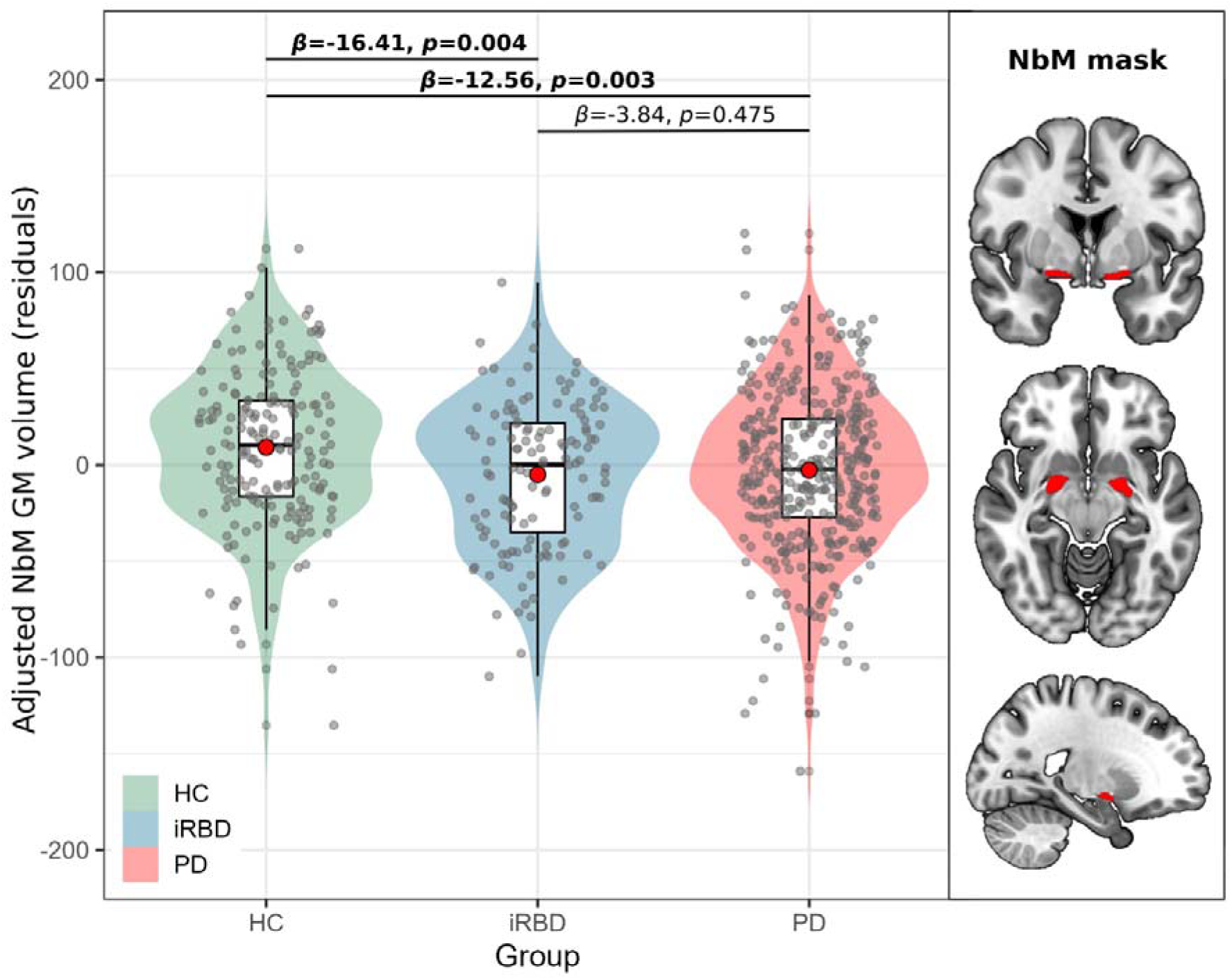
Between-group differences in grey matter volume of the NbM. Significantly lower NbM grey matter volume was found in the iRBD and PD groups compared to controls. No significant differences were observed between the iRBD and PD groups. Red circles within boxplots represent group mean. Right side: bilateral NbM mask in red. Group-differences with *p*<0.05 are highlighted with bold. P-values are FDR-corrected.

### Cognitive assessment

To assess cognitive function, we used the Montreal Cognitive Assessment (MoCA) which was available for both Oxford Discovery and PPMI, and higher-order cognitive testing that was only available in PPMI participants. Raw MoCA scores were adjusted for education years as recommended^37^.

In the PPMI cohort, cognitive tests previously shown to be associated to disease with cholinergic deficits were selected, namely visuospatial skills, attention, executive functions, and memory^5,11–13^. Visuospatial function was evaluated using the Benton Judgment of Line Orientation test, verbal memory was assessed using total immediate recall (encoding) and delayed recall (retrieval) scores of the Hopkins Verbal Learning Test–Revised, executive functions were assessed by means of the Letter-Number Sequencing test and the Semantic Verbal Fluency test, and attention by the Symbol Digit Modalities Test (SDMT)^29,38^. Since all these tests were only available as part of the PPMI study, only the participants from PPMI were analysed in this context. In order to create a higher-order cognition composite score we first fitted linear regression for each test score controlling for age, sex, and years of education on 279 healthy controls from the PPMI database with cognitive data at baseline visit (due to a notable ceiling effect in the Benton Judgement of Line Orientation test scores, we used a Tobit model for this test using the *‘VGAM’* package in R)^39^. We then applied the model to obtain expected test scores given age, gender and years of education in the PD and iRBD groups. Next, we computed the difference between true score and expected score and z-scored the differences. Lastly, we averaged all single z-scores to obtain the final higher-order cognition composite score^40,41^.

### Disease severity measures

We used the total score of The MDS-UPDRS part 1 and the total score of the MDS-UPDRS part 3, respectively, to assess the relationship between the grey matter volume of the NbM and the extent of non-motor and motor symptoms in iRBD and PD patients^42^. In iRBD participants only, we calculated the probability of prodromal PD^43^ based on the recent MDS Research Criteria for Prodromal Parkinson’s Disease^44^. For that, we used only risk and prodromal markers available for both the ODC and PPMI cohorts, namely, age group, sex, subthreshold parkinsonism (UPDRS-3 excluding active and postural tremor), olfactory loss, constipation, first degree relative with PD, excessive daytime somnolence, orthostatic hypotension, urinary dysfunction, and depression. In addition, we ran two models, with and without taking PSG-proven RBD diagnosis into account in the calculation of the prodromal probability, as all RBD participants in the current study were PSG-diagnosed with RBD.

### Conversion to dementia in PD

Dementia criteria for the ODC participants were based on a combination of the MoCA score and the presence of cognitive-related functional impairment in the ‘Cognitive impairment’ subsection of the UPDRS-1^7,45,46^. Specifically, we used a combination of a MoCA score ≤21 and a UPDRS-1 cognitive impairment score ≥2 (i.e., clinically evident cognitive dysfunction and evidence of some degree of interference with everyday-life functioning)^8,45–47^. In the PPMI study, cognitive diagnosis, i.e., normal cognition, MCI, or dementia is conducted at each visit by the site investigator, based on assessment of cognitive change compared with pre-Parkinson’s state, impairment in cognitive abilities, and resulting functional impairment^7,48^. For the analyses examining conversion to dementia in PD in the current study (detailed in the *Statistical analysis* section below), we ran two different models in which PPMI converters to dementia were defined based on either the investigator assignment of cognitive diagnosis or the MoCA-UPDRS-1 combination (in accordance with the ODC participants). We excluded participants who were classified with dementia (either by the site investigator or the MoCA-UPDRS criteria) at one visit but then reverted to a non-dementia state (based on those criteria) on a later visit, indicating possible previous misclassification.

### Phenoconversion in iRBD

In the current study we have focused on the two major types of phenoconversion among iRBD patients (i.e., to either PD or DLB) and excluded participants who were diagnosed with other neurodegenerative disorder (e.g., one participant with multiple systems atrophy (MSA)). Diagnosis of iRBD to PD or DLB phenoconversion in the ODC was performed using standard diagnostic criteria^49,50^ applied by trained neurologists assessing each patient longitudinally with a structure series of assessments and examination, with carer report as appropriate. Phenoconversion in the PPMI dataset was defined as individuals with iRBD who were given a diagnosis of either PD or DLB on a follow-up visit.

### Statistical analysis

Statistical analyses and visualizations were performed using R version 4.4.0 (https://www.r-project.org/). To assess whether NbM volume differed between PD, iRBD, and controls, we first summed the right and left NbM volumes to a single bilateral measure (i.e., NbM volume) due to the high correlation between the volumes of the two sides (r=0.83). We used linear mixed model analyses using the *‘lme4’* package implemented in R to examine group differences in the measures of interests and continuous associations with NbM volume. In addition, as a complementary analysis, we also divided the PD group into those with and without possible concomitant RBD, as PD with RBD is considered a more clinically aggressive subtype^28^. To this end, we used the RBD Screening Questionnaire (RBDSQ) to assess RBD symptoms^51^. This questionnaire contains 13 items that measure the history of occurrence of dreams, dream-related behaviours, consequence of the behaviours, and other nervous system diseases with a yes or no response. In this study, we used a cutoff of ≥6 to classify patients to either PD with possible RBD (PD + pRBD) or without (PD - pRBD)^28,52,53^. Three patients with PD were excluded because they did not have an RBDSQ available. P-values for all pairwise differences were corrected with the False Discovery Rate (FDR) method^54,55^.

For each analysis, we first ran a full model with by-site random intercept and random slope for the explanatory variable of interest. Whenever the fitted mixed model resulted in a singular model due to zero variance of the random slope, we reduced the model to a random intercept only model. Whenever a random intercept model resulted in a singular model (due to zero variance in the random effect) we ran a multivariable linear regression model. Since a substantial amount of the iRBD patients in the current study presented with >99% calculated probability of prodromal PD when including PSG-diagnosis in the calculation of the prodromal probability, we ran a Tobit regression to evaluate its relationship with NbM volume^39^. When not including PSG-diagnosis in the calculation, the prodromal probability followed a positive skewed distribution. Hence, we used an inverse Gaussian regression, which is a form of the generalized linear model when the response variable is continuous and positively skewed^56^. In all models we controlled for age, sex, years of education, and disease duration, i.e., time from clinical diagnosis (PSG-iRBD or PD) to baseline visit in months) as potential confound variables. We included TIV and IQR as covariates in all models in which NbM volume was used as independent or dependent variable. The models’ coefficients (with their confidence intervals) reported throughout the text represent the average change in the raw outcome variable that is associated with a 1 standard deviation change in the predictor variable.

To test the association between baseline NbM volume and the risk of conversion to dementia among PD patients or the risk of phenoconversion among the iRBD patients, we performed Cox proportional hazards regression analyses using the *‘survival’* package in R. In all survival models we used a frailty term for site as a grouping variable. In the model estimating the risk for dementia in PD we also included years of education and sex as additional covariates. We did not add sex as a covariate to the iRBD model due to the very high proportion of male patients in this group (∼90% among all iRBD patients and all phenoconverters in the current study). Due to the low number of iRBD participants phenoconverting to DLB (n=8), phenoconversion in the models was defined as either DLB or PD diagnosis, i.e., collapsed across these conditions. Time of follow-up in the models was defined as the time interval in months between baseline visit and last visit for patients who did not convert and between baseline visit and date of conversion diagnosis among converted patients. Only patients who had at least one additional clinical evaluation after their baseline visit were included in these analyses. Cases were censored when dementia criteria fulfilled/phenoconversion were clinically diagnosed or at their last visit. Schoenfeld residuals method was used to verify that the assumption of proportional hazards was not violated.

## Results

### Demographics

Participants demographics are presented in Table 1. Extended table with PD +/- pRBD groups is presented in **Supplementary Table 2**.

### Group differences in NbM grey matter volume

When we compared the three groups, we found lower NbM grey matter volume in both the iRBD group (β=-16.41 [95% confidence interval (CI) −26.63, −6.19], *pFDR*=0.004) and PD group (β=-12.56 [95% CI −19.96, −5.17], *pFDR*=0.003) compared to controls, controlling for age, sex, years of education, TIV, and IQR (**Figure 1),** while no such difference was found between iRBD and PD groups (*pFDR*=0.48). When dividng the PD group based on the RBDSQ score, both PD + pRBD (β=-17.79 [95% CI −27.92, −7.66], *pFDR*=0.003) and PD – pRBD (β=-10.34 [95% CI −18.16, −2.52], *pFDR*=0.003) demonstrated lower NbM volume compared to controls, with no difference between the iRBD and PD groups (*pFDR*>0.19 for all pairwise comparisons) (see **Supplementary Figure 1**).

### Association between NbM grey matter volume and cognitive function

#### General cognitive function - MoCA

First, we found no evidence of differences in general cognitive function, as measured with the MoCA, for iRBD and PD (β=-0.16 [95% CI −0.88, 0.56], *p*=0.67), controlling for age, sex and years of education. Similarly, the association between NbM grey matter volume and overall MoCA scores in either the iRBD group (β=-0.20 [95% CI −0.72, 0.32], *p*=0.45) or the PD group (β=0.11 [95% CI −0.17, 0.39], *p*=0.43) (**Figure 2a**), was consistent with chance (controlling for age, sex, years of education, disease duration, TIV, and IQR). Similar metrics were observed in the PD group when controlling for possible RBD level (β=0.10 [95% CI - 0.18, 0.38], *p*=0.48).

**Figure 2.**
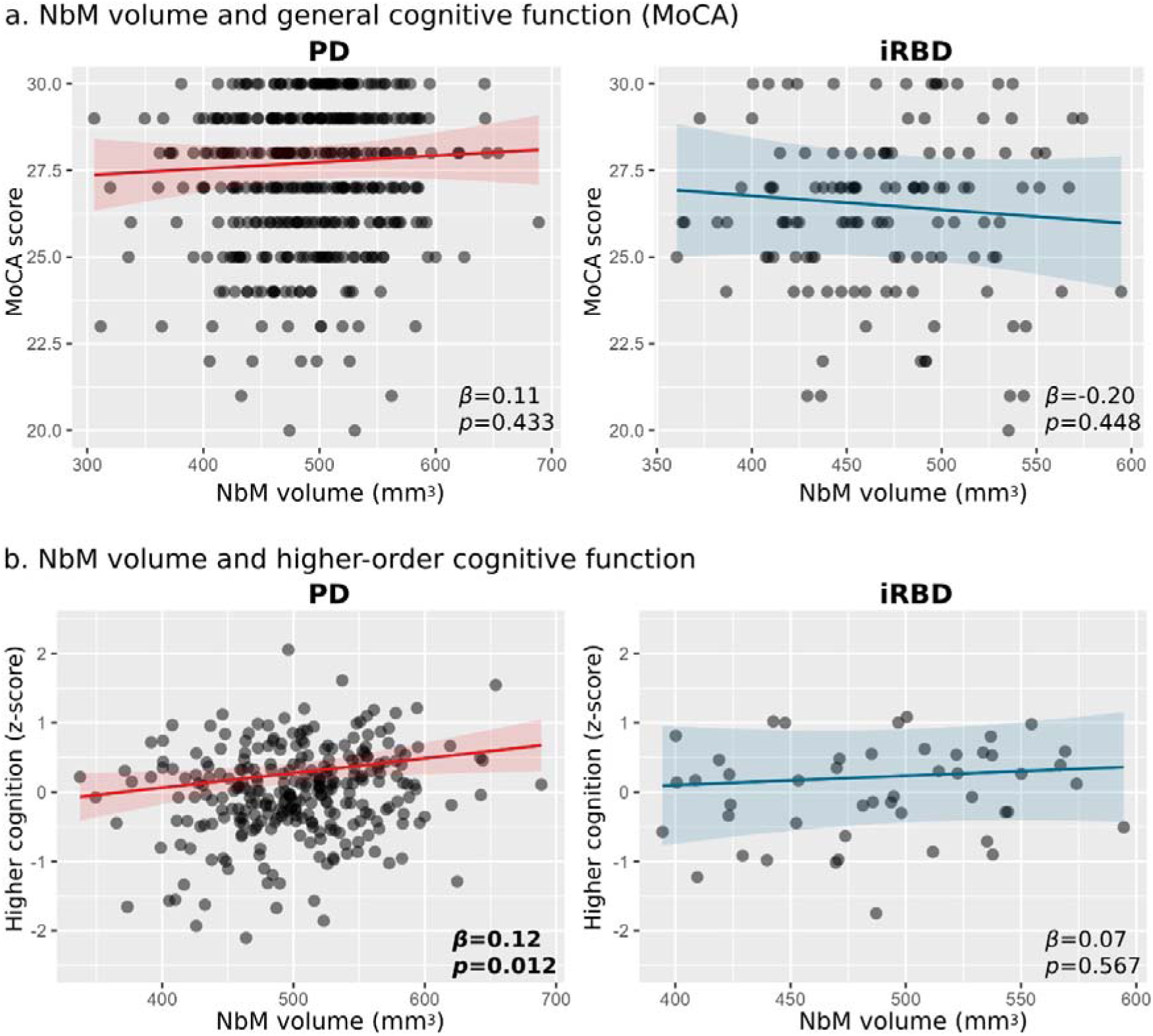
NbM structural deficit and cognitive function in prodromal and early Lewy body disease. (a) NbM volume was not statistically associated with general cognitive performance as measured by the MoCA test in either PD (left panel) or iRBD (right panel). (b) Higher NbM volume was associated with better higher-order cognitive function in PD patients (left panel) but not in iRBD (right panel). Parameter estimates in scatter plots represent standardized model coefficients for NbM volume. Associations with *p*<0.05 are highlighted with bold.

#### Higher-order cognitive function

Since only available in the PPMI dataset, we examined whether higher-order cognitive function is related to NbM degeneration in the PD (n=288) and iRBD (n=48) participants in PPMI, as previously suggset in Lewy body disease^5^. We did not observe significant difference between the iRBD and PD patients with regard to their average composite higher-order Z-score performance, when controlling for age, sex, and years of education (β=0.03 [95% CI - 0.21, 0.26], *p*=0.84). Higher NbM grey matter volume was found to be associated with higher-order cognitive function in the PD group (β=0.12 [95% CI 0.03, 0.22], *p*=0.01), but not in the iRBD patients (β=0.07 [95% CI −0.18, 0.32], *p*=0.57), though the 95% confidence interval for the latter was wide and overlapped with the PD group effect estimate (**Figure 2b**). Similar metrics were observed in the PD group when controlling for possible RBD level (β=0.11 [95% CI 0.02, 0.21], *p*=0.02).

### Association between NbM grey matter volume and motor and non-motor clinical symptoms

#### UPDRS-1 total score

When comparing the groups with regard to the non-motor part of the UPDRS-1, we found iRBD patients to have significantly higher UPDRS-1 total score compared to the PD group, when controlling for age, sex, and education (β=1.59 [95% CI 0.58, 2.60], *p*=0.002). This difference was even higher when UPDRS-3 score was added and controlled for in the model (β=2.93 [95% CI 1.57, 4.29], *p*<0.001). Higher NbM was associated with lower UPDRS-1 total score in PD patients (β=-0.66 [95% CI −1.25, −0.08], *p=*0.03). Association in the same direction, though consistent with chance probability, was observed in the iRBD group (β=0.29 [95% CI −0.87, 1.45], *p*=0.62) (**Figure 3a**). Similar metrics were observed in the PD group when controlling for possible RBD level (β=-0.63 [95% CI −1.21, −0.04], *p*=0.04).

**Figure 3.**
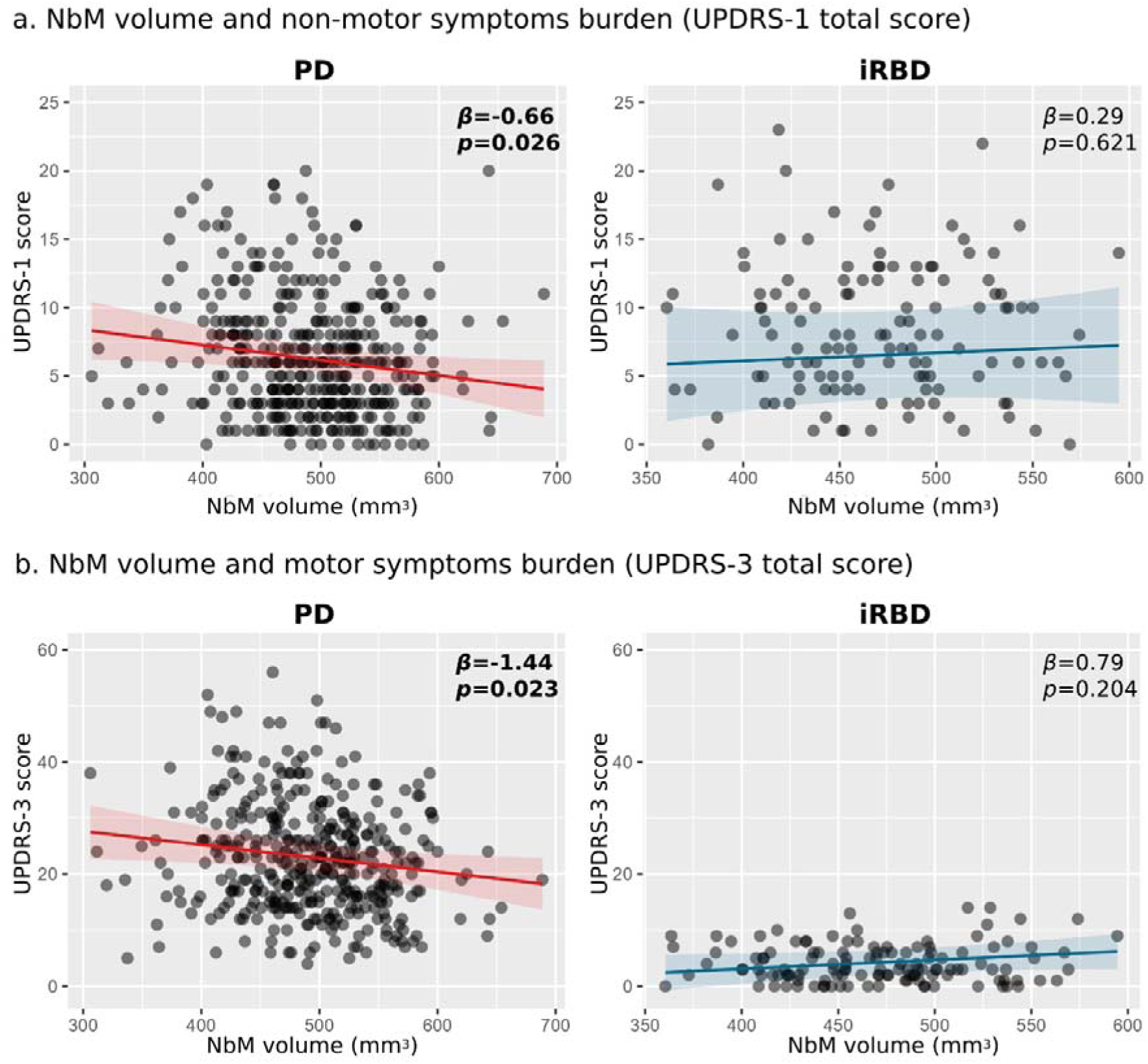
NbM gray matter volume and measures of disease severity in prodromal and early Lewy body disease. (a) Higher NbM volume was significantly associated with decreased non-motor symptoms severity/lower UPDRS-1 total score in PD patients (left panel) but not among iRBD (right panel). (b) Higher NbM volume was associated with decreased motor symptoms severity/lower UPDRS-3 total score in PD patients (left panel) but not among iRBD (right panel). Y-axes ranges were compared for both groups’ plots to appreciate the range of each measurement in each group compared to the other. Parameter estimates in scatter plots represent standardized model coefficients for NbM volume. Associations with *p*<0.05 are highlighted with bold.

#### UPDRS-3 total score

As expected, iRBD patients demonstrated significantly lower UPDRS-3 total score compared to PD patients (β=-19.81 [95% CI 17.98, 21.63], *p*<0.001). Higher NbM volume was associated with lower UPDRS-3 score in PD patients (β=-1.44 [95% CI −2.68, −0.20], *p*=0.02). In the iRBD group higher NbM volume was not statistically associated with UPDRS-3 score (β=0.79 [95% CI −0.17, 1.76], *p*=0.20) (**Figure 3b**). Similar metrics were observed in the PD group when controlling for possible RBD level (β=-1.38 [95% CI −2.62, −0.14], *p*=0.03).

#### Prodromal probability of PD

NbM volume was not statistically associated with prodromal probability of PD in iRBD, either when PSG-diagnosis was included in the calculation (β=-0.04 [95% CI −0.09, 0.01], *p*=0.09) or not (β=0.56 [95% CI −0.67, 1.79], *p*=0.37), when controlling for TIV, IQR, years of education, disease duration, and site as a fixed effect. We did not control for factors such as age, sex, or other disease severity markers as they were already used in the calculation of the prodromal probability score.

### Association between NbM grey matter volume and risk of conversion

#### Converting to PD dementia among PD patients

A total of 24 patients with PD from both PPMI and ODC converted to PD dementia during a follow-up period of 69.04±48.8 months, when using the site investigator diagnosis in PPMI. Using a multivariable Cox regression, we found increase of 1 standard deviation of NbM grey matter volume to be associated with ∼60% reduced risk of converting to dementia among the PD patients (HR=0.399 [95% CI 0.216, 0.738], *p*=0.003, **Table 2**, **Figure 4a**), when controlling for age at baseline, sex, disease duration at baseline, years of education, TIV, and IQR. Similar metrics, i.e., ∼62% reduced risk, were found when using the MoCA-UPDRS-1 criteria for all participants (HR=0.381 [95% CI 0.205, 0.707], *p*=0.002), which resulted in 23 dementia converters following a follow-up period of 69.27±48.8 months (**Table 2**, **Figure 4a**). Similar metrics, i.e., ∼61-63% reduced risk, were further observed when controlling for possible RBD level, as well as baseline MoCA, UPDRS-3 and UPDRS-1 total scores in both models (site investigator: HR=0.387 [95% CI 0.207, 0.723], *p*=0.003; MoCA-UPDRS-1: HR=0.366 [95% CI 0.199, 0.674], *p*=0.001, **Table 2**)

**Figure 4.**
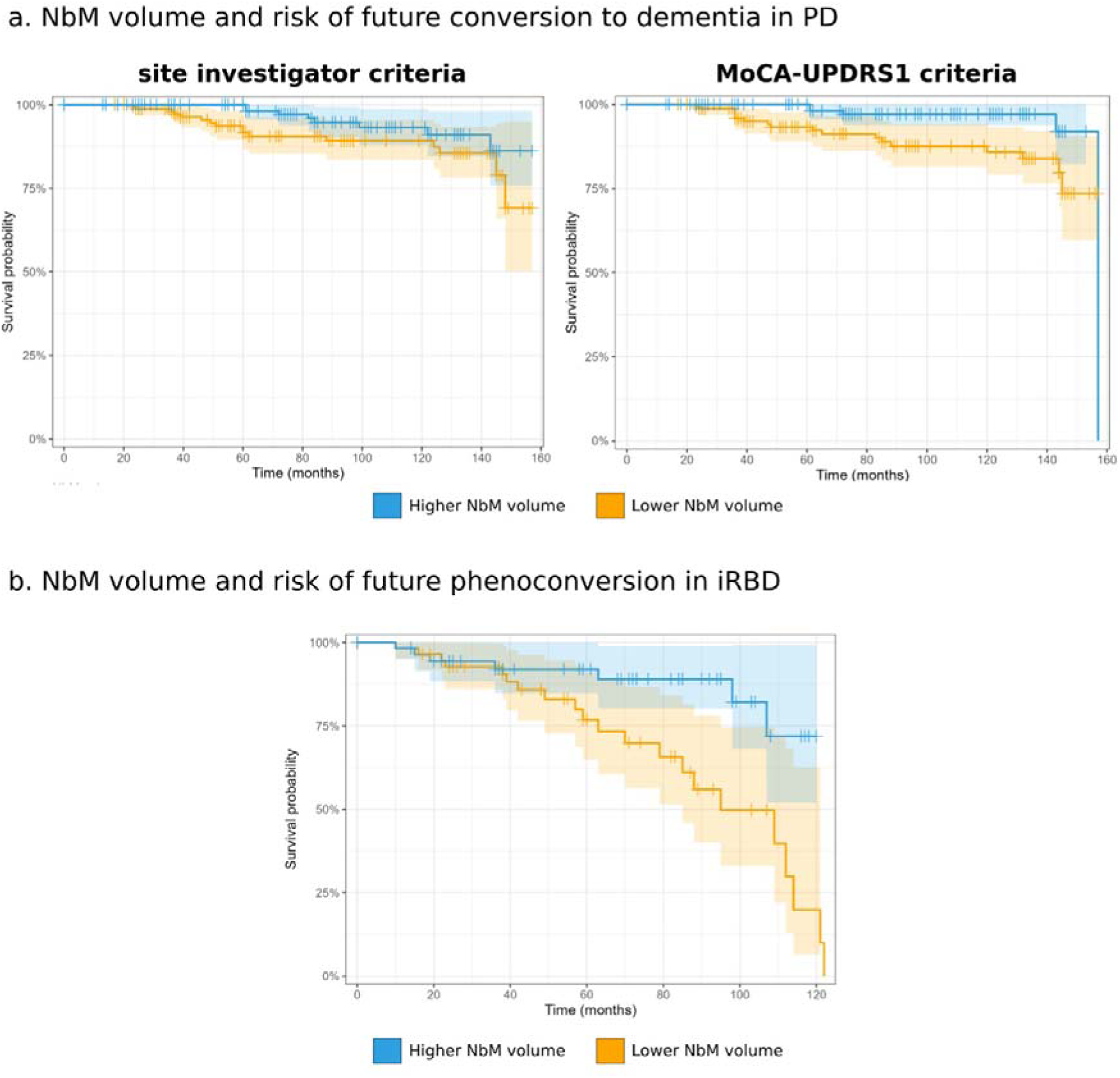
NbM structural deficit and risk of future conversion in prodromal and early PD. Survival probabilities of converting to dementia in PD (a) and phenoconversion in iRBD (b) given higher or lower NbM volume at baseline visit based on median split of adjusted NbM volume (i.e., residuals) for visualization purposes.

**Table 2.**
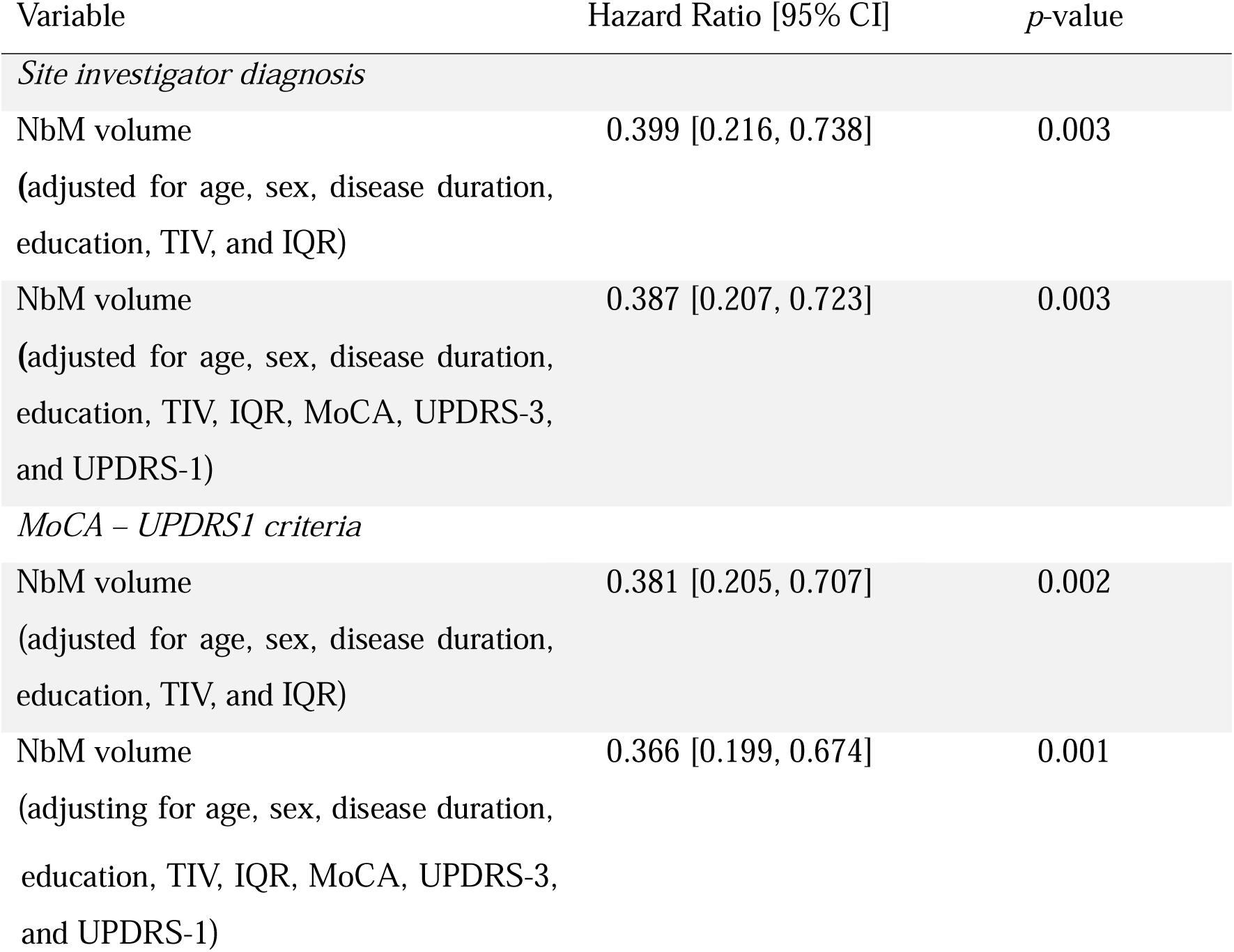
Survival analysis of NbM volume and risk of dementia conversion in PD patients. Hazard Ratio coefficients are per 1 standard deviation of NbM volume.

#### Phenoconversion in iRBD

During a follow-up period of 55.0±35.9 months, 28 patients with iRBD from both PPMI and ODC phenoconverted to either PD (n=20) or DLB (n=8) during the follow-up period. We found 1 standard deviation increase in NbM grey matter volume at baseline to be associated with ∼51% reduced risk of future phenoconversion among iRBD patients when controlling for age at baseline, disease duration at baseline, years of education, TIV, and IQR (HR=0.484 [95% CI 0.269, 0.871], *p*=0.015, **Table 3**, **Figure 4b**). A ∼55% reduced risk, were found when UPDRS-3 and UPDRS-1 total scores were also added and controlled for in the model (HR=0.444 [95% CI 0.247, 0.80], *p*=0.007, **Table 3**.

**Table 3.** Survival analysis of NbM volume and risk of phenoconversion in iRBD patients. Hazard Ratio coefficients are per 1 standard deviation of NbM volume.

| Variable | Hazard Ratio [95% CI] | *p*-value |
| --- | --- | --- |
| NbM volume<br>(adjusted for age, disease duration,<br>education, TIV, and IQR) | 0.484 [0.269, 0.871] | 0.015 |
| NbM volume<br>(adjusted for age, disease duration,<br>education, TIV, IQR, UPDRS-3, and<br>UPDRS-1) | 0.444 [0.247, 0.800] | 0.006 |

## Discussion

In this comprehensive multi-cohort study, we demonstrate that nucleus basalis of Meynert volume is significantly reduced in both early manifest PD and in iRBD, a key disease-prodromal state of synucleinopathies. Critically, we found that NbM volume holds predictive value for disease progression, with lower volumes associated with increased risk of conversion to dementia in PD and phenoconversion in iRBD patients. These findings remained robust even after controlling for potential confounding factors including age, disease duration, and education level, suggesting that NbM volume could serve as an independent biomarker for risk-stratifying patients in clinical trials of disease-modifying therapies.

The relationship between NbM integrity and disease manifestation showed distinct patterns across disease stages. In early PD, reduced NbM volume was significantly associated with poorer higher-order cognitive function, increased motor symptom severity, and greater non-motor burden. This constellation of associations suggests that NbM degeneration may represent a key substrate of clinical heterogeneity in early PD. Interestingly, while iRBD patients showed comparable magnitude of NbM volume reduction to PD patients when compared to controls, a relationship between NbM integrity and clinical features was not evident at this prodromal stage, despite the predictive value for future phenoconversion. This dissociation suggests that compensatory mechanisms may initially mask the clinical impact of cholinergic dysfunction in prodromal disease stages, with subsequent failure of these mechanisms potentially contributing to clinical conversion.

NbM degeneration in prodromal and manifest Lewy body patients has been reported on autopsy, showing significant cell loss in the NbM in PD patients with and without dementia^16^. However, previous works utilizing MRI have demonstrated inconsistent results regarding MRI-derived NbM degeneration in PD. For example, Schulz et al. (2018) found no difference between NbM grey matter volume in PD and controls^57^, Ray et al. (2018) only found lower volume in the posterior NbM (but not the entire NbM) in PD with mild cognitive impairment (but not without cognitive decline) compared to controls^58^, while Schumacher et al. (2023) showed significant NbM volume loss in PD^59^.

Here, in addition to the group-level differences in NbM volume that were observed in PD, this measure was also found to have value with regard to subject-level associations with baseline and longitudinal clinical-behavioural measures. These results complement previous findings by which lower grey matter volume of the NbM was found to be predictive of longitudinal cognitive decline in PD, but crucially, we were able to demonstrate this association based on two cohorts and longer follow-up periods^57^. While up to >80% of PD patients are eventually anticipated to develop dementia, there is a large variability in the time interval between initial PD diagnosis and the diagnosis of dementia among those patients^5^. This makes the identification of factors and biomarkers with a predictive value for time to conversion crucial for the design of future interventional studies, but also helps decision making for patients, families/carers, and healthcare professionals. In addition, a relatively simple measurement such as the grey matter volume of the NbM derived from non-invasive structural MRI, could be used to enrich future studies aiming to prevent or delay the onset of dementia in PD, by identifying patients with higher risk of near-term conversion.

In the current study, significantly lower NbM volume was also observed in patients with iRBD compared to controls, but no difference between iRBD and PD was found. This finding is consistent with the only previous study to our knowledge that examined NbM grey matter in iRBD. A smaller study by Tan et al. (2023) found reduced grey matter density in the Ch4 region, but not in Ch1-3, compared to controls^60^. In comparison to manifest Lewy body disorders, neuropathological evidence on the structural and functional correlates of the BFCS in iRBD is scant. Our findings suggest that degeneration of the NbM is evident already at this prodromal subtype of Lewy body disorder. In addition, the current study is the first to our knowledge presenting evidence for a relationship between NbM degeneration and future risk of phenoconversion in iRBD. However, in the current study we could not make a firm conclusion about the added value of NbM volume in differentiating between the phenoconversion types (i.e., PD vs. DLB). This is due to the small number of participants phenoconverting to DLB, which could result from several reasons such as insufficient follow-up time interval, self-selection bias, and the overall larger proportion of iRBD converting to PD compared to DLB^26^.

While higher NbM volume was associated with better performance on higher-order cognitive tests in PD, no such effect was found in iRBD. NbM volume was also not associated with MoCA performance in either iRBD or PD. These findings complement two previous studies which also did not find associations between cholinergic markers in the brain, namely acetyl-cholinesterase (AChE) activity levels, and the Mini Mental State Examination (MMSE), MoCA, or the MoCA subitems for visuospatial functions at baseline^61^ or between the extent of reduction in measured AChE over 3 years and changes in performance on those tests^62^. This indicates a potential ceiling effect in the MoCA score, which may limit the clinically meaningful performance range among patients at the top end of its range. This is inherent to its design, which aims to identify manifest cognitive dysfunction, rather than detecting subtle prodromal change, although it scales better at the upper range of performance than the MMSE^63^.

What then may underly the lack of association in iRBD, despite the role of ACh in clinical cognitive decline? Isolated RBD appears to represent a more aggressive subtype of PD compared to idiopathic PD without premotor RBD^23,24^. iRBD patients already suffer with a high rate of autonomic, neuropsychiatric, and olfactory symptoms^26^, which is also evident in the current study by the higher UPDRS-1 score observed compared to the PD group. This suggests that multiple neural pathological processes play a role in iRBD, potentially affecting different neuromodulatory systems in the brain, which in turn contributing to different clinical and behavioural expressions. NbM integrity is only one aspect of this multidimensional process. Future studies investigating the integration of several neuromodulatory systems (i.e., cholinergic, dopaminergic, noradrenergic, and serotonergic) and their interactions with clinical symptoms and phenoconversion would be valuable in unravelling some of this complexity.

The basal forebrain cholinergic system is composed of not only the grey matter of the Ch1-4 regions, but also the white matter tracts originating from them, and the presynaptic terminals that target cortical/subcortical neurons. It has been proposed that the degeneration of NbM neurons precedes and triggers the loss of their axons that are directed to the cortex^64^. This would indicate a sequential process, contributing to the overall expression of cognitive and clinical features in more advanced stages of disease. While a relationship between NbM volume and PET measures of cortical cholinergic activity was previously reported in PD patients^59^, previous studies showed inconsistent results in terms of PET markers of cholinergic deficits in patients with iRBD. While Bedard and colleagues found higher vesicular acetylcholine transporter levels, a proxy for presynaptic cholinergic integrity, in the cortex and brainstem^65^, Gersel Stokholm and colleagues observed reduced cortical AChE levels^61^, suggesting a complex interplay of pathology and compensatory response. This may suggest that during early stages of Lewy body disease, compensatory mechanisms may act in some of the patients to preserve levels of cholinergic activity despite the structural deficits, similar to what has been proposed to take place in early stages of PD with regard to nigro-striatal adaptations^66^. It is possible that only later at the course of the neuropathological process, when those mechanisms are no longer effective in compensating for the structural loss, the association between cholinergic deficits and clinical/cognitive features may become clinically evident, as in the case of PD and DLB patients. Furthermore, impaired neurotransmitter release from cholinergic terminals may also contribute to the inconsistent observations with ACh PET studies, similar to what has been reported to occur in dopaminergic neurons in PD^67^.

### Limitations

NbM degeneration and cell loss have been suggested to occur contemporaneously with nigral pathology during the course of PD at the microscopic level^14^. The MRI-derived grey matter contrast, which is used to estimate macroscopic NbM volume, is not a direct measure of cellular density or integrity. Several neuronal and non-neuronal tissue properties may be involved, and reflect different intracellular and extracellular processes^68^. Theses may differ between patients, and at least partially underlie the differences observed between cohorts and studies. In addition, the NbM volume represents one component of the vast cholinergic network between the basal forebrain and the cortex, and other parts of this system may independently and additively contribute to its overall (dys)function. For example, in a recent study in PD patients combining MRI with cholinergic PET, only a weak-to-moderate relationship (r=0.29) was found between the posterior basal forebrain volume (mainly corresponding to the NbM) and cortical acetylcholinesterase activity^59^. Furthermore, in the same study, the two cholinergic markers were found to contribute differently to different cognitive functions. Also, as previously demonstrated, the specific NbM region of interest chosen and the method to define it in the imaging volume will affect study-specific results^36^. As best practice, we followed established selection criteria for realistic NbM volume estimation^36^. Differences in analysis pipelines^69,70^, along with differences in sample sizes (and therefore statistical power), are also likely to contribute to this discrepancy. The sample sizes for PD and iRBD in the current study were substantially different (393 vs. 128, respectively), which affected within-group statistical power, especially in the case of the iRBD participants. In the current study we did not directly examine group-based interaction effects for the different clinical and behavioural metrics, due to the substantial differences in groups sizes and given that it has been suggested that substantially larger sample sizes are required for the identification of interaction effects compared to main effects^71^.

### Conclusions

NbM structural integrity provides valuable prognostic information across of Lewy body associated disease, from prodromal to early clinical stages. In established PD, NbM volume not only correlates with current cognitive and clinical status but also predicts future dementia risk, suggesting its utility as a biomarker for clinical trials targeting cognitive decline. While NbM degeneration is evident in iRBD before clinical parkinsonism or dementia emerge, its relationship to symptoms appears more complex, possibly reflecting early compensatory mechanisms. The predictive value of NbM volume for phenoconversion in iRBD highlights its potential as an early marker of neurodegeneration, particularly valuable for identifying high-risk individuals for neuroprotective interventions. Future research should focus on understanding the temporal dynamics of cholinergic system deterioration and its interaction with other neurotransmitter systems, which may reveal new therapeutic windows for intervention before irreversible clinical progression occurs.

## Data Availability

PPMI data are freely accessible to researchers by signing a Data Use Agreement on the PPMI website. (https://www.ppmi-info.org). ODC data are available upon reasonable request. Qualified investigators seeking access to de-identified participant data relating to the ODC may submit their request by means of a formal application to the Oxford Parkinson’s Research Centre (OPDC) Data Access Committee. The application form, protocol, and terms and conditions may be found at <u>opdc.medsci.ox.ac.uk/external-collaborations</u>.

## Supplementary Materials

**Supplementary Table 1.** Contribution of each cohort to the study groups’ participants.

| Cohort | PD<br>(N = 393) | iRBD<br>(N = 128) | HC<br>(N = 186) |
| --- | --- | --- | --- |
| ODC | 105 | 80 | 59 |
| PPMI | 288 | 48 | 127 |

**Supplementary Table 2.** demographic, cognitive, and clinical characteristics of study participants (mean±SD and proportions are presented for continuous and categorical variables, respectively).

| Characteristic | PD<br>(N = 393) | PD + pRBD<br>(N = 104) | PD - pRBD<br>(N = 286) | iRBD<br>(N = 128) | HC<br>(N = 186) |
| --- | --- | --- | --- | --- | --- |
| Age (years) | 63.5 (9.54) | 64.7 (9.06) | 63.0 (9.63) | 67.2<br>(7.23) | 62.6 (10.91) |
| Sex |  |  |  |  |  |
| Female | 149 (38%) | 25 (24%) | 121 (42%) | 13 (10%) | 68 (37%) |
| Male | 244 (62%) | 79 (76%) | 165 (58%) | 115<br>(90%) | 118 (63%) |
| Education<br>(years) | 15.7 (3.12) | 15.6 (3.03) | 15.8 (3.16) | 14.4<br>(3.40) | 16.0 (3.09) |
| MoCA<br>score | 27.3 (2.10) | 27.1 (2.23) | 27.3 (2.06) | 26.3<br>(2.30) | 27.9 (2.00) |
| Higher<br>order<br>cognition<br>(z-score) | -0.01<br>(0.64)<br>(N = 282) | -0.19 (0.66)<br>(N = 209) | 0.06 (0.63)<br>(N = 72) | 0.00<br>(0.68)<br>(N = 48) | - |
| UPDRS-1<br>total score | 6.4 (4.30) | 6.9 (4.37) | 6.2 (4.28) | 8.5<br>(5.13) |  |
| UPDRS-3<br>total score | 22.8 (9.68) | 24.5 (11.18) | 22.3 (9.04) | 4.1<br>(3.37) | - |
| Disease<br>duration<br>(months) | 9.1 (8.96) | 9.1 (8.93) | 9.0 (8.88) | 27.3<br>(44.37) | - |
| TIV (ml) | 1,531.2<br>(152.86) | 1,550.7<br>(127.59) | 1,525.7<br>(160.43) | 1,541.1<br>(125.34) | 1,495.9<br>(146.81) |
| IQR | 80.7 (3.66) | 80.6 (3.82) | 80.7 (3.61) | 81.6<br>(3.95) | 80.3 (3.74) |

**Supplementary Figure 1.**
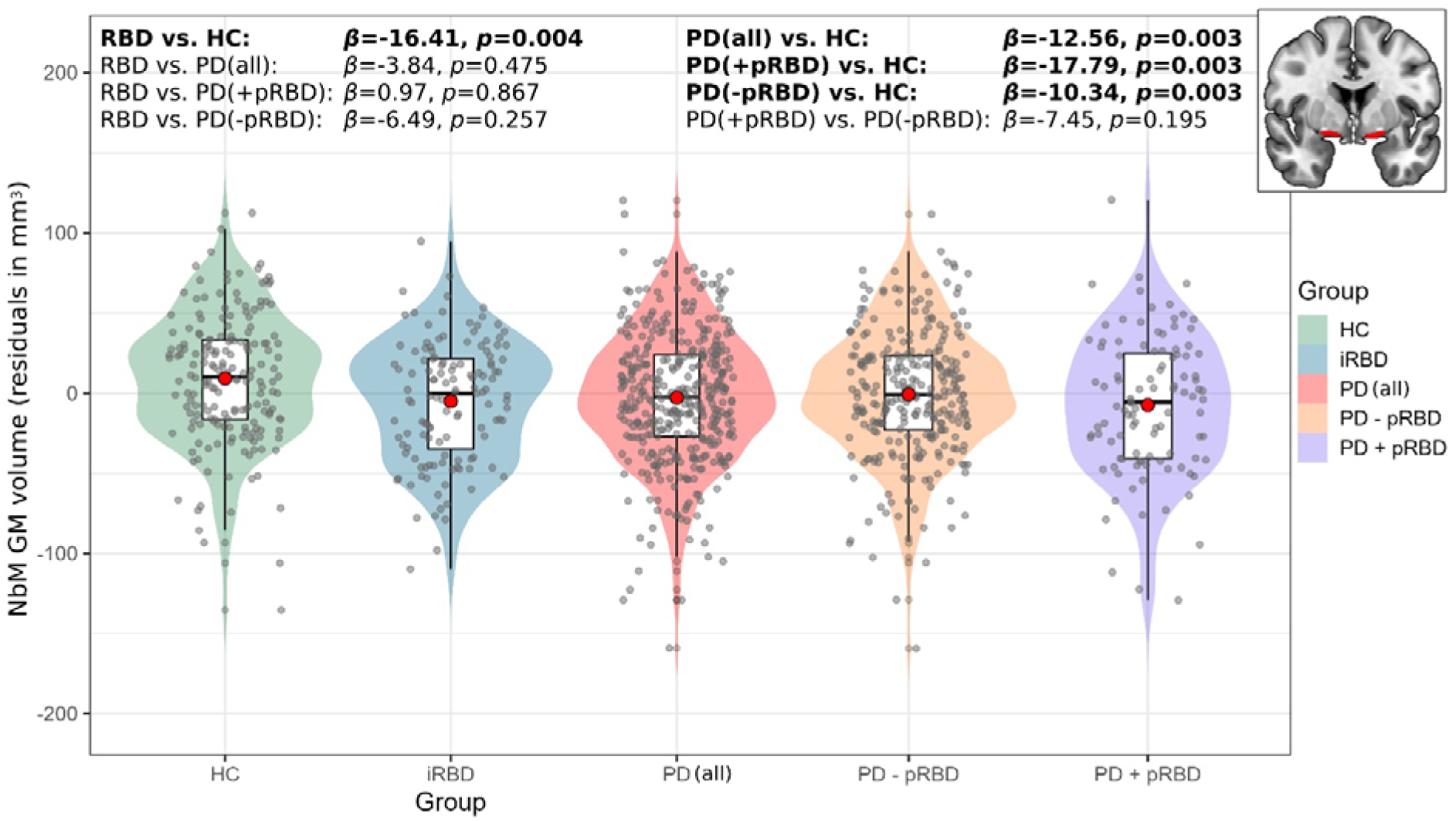
Between-group differences in grey matter volume of the NbM. Significantly lower NbM grey matter volume was found between iRBD and all PD groups compared to controls. No significant differences were observed between iRBD and PD groups. Red circles within boxplots represent group mean. Upper right corner: bilateral NbM mask in red. Group-differences with *p*<0.05 are highlighted with bold. P-values are FDR-corrected. PD + pRBD = PD with possible RBD; PD - pRBD = PD without possible RBD

